# SARS-CoV-2 antibodies remain detectable 12 months after infection and antibody magnitude is associated with age and COVID-19 severity

**DOI:** 10.1101/2021.04.27.21256207

**Authors:** Eric D. Laing, Nusrat J. Epsi, Stephanie A. Richard, Emily C. Samuels, Wei Wang, Russell Vassell, Daniel F. Ewing, Rachel Herrup, Spencer L. Sterling, David A Lindholm, Eugene V. Millar, Ryan C. Maves, Derek T. Larson, Rhonda E. Colombo, Sharon Chi, Cristian Madar, Tahaniyat Lalani, Anuradha Ganesan, Anthony Fries, Christopher J. Colombo, Katrin Mende, Mark P. Simons, Kevin L. Schully, Carol D. Weiss, David R. Tribble, Brian K. Agan, Simon D. Pollett, Christopher C. Broder, Timothy H. Burgess, for the EPICC Study team

## Abstract

**Importance:** The persistence of SARS-CoV-2 antibodies may be a predictive correlate of protection for both natural infections and vaccinations. Identifying predictors of robust antibody responses is important to evaluate the risk of re-infection / vaccine failure and may be translatable to vaccine effectiveness.

**Objective:** To 1) determine the durability of anti-SARS-CoV-2 IgG and neutralizing antibodies in subjects who experienced mild and moderate to severe COVID-19, and 2) to evaluate the correlation of age and IgG responses to both endemic human seasonal coronaviruses (HCoVs) and SARS-CoV-2 according to infection outcome.

**Design:** Longitudinal serum samples were collected from PCR-confirmed SARS-CoV-2 positive participants (U.S. active duty service members, dependents and military retirees, including a range of ages and demographics) who sought medical treatment at seven U.S. military hospitals from March 2020 to March 2021 and enrolled in a prospective observational cohort study.

**Results:** We observed SARS-CoV-2 seropositivity in 100% of inpatients followed for six months (58/58) to one year (8/8), while we observed seroreversion in 5% (9/192) of outpatients six to ten months after symptom onset, and 18% (2/11) of outpatients followed for one year. Both outpatient and inpatient anti-SARS-CoV-2 binding-IgG responses had a half-life (*T*_1/2_) of >1000 days post-symptom onset. The magnitude of neutralizing antibodies (geometric mean titer, inpatients: 378 [246-580, 95% CI] versus outpatients: 83 [59-116, 95% CI]) and durability (inpatients: 65 [43-98, 95% CI] versus outpatients: 33 [26-40, 95% CI]) were associated with COVID-19 severity. Older age was a positive correlate with both higher IgG binding and neutralizing antibody levels when controlling for COVID-19 hospitalization status. We found no significant relationships between HCoV antibody responses and COVID-19 clinical outcomes, or the development of SARS-CoV-2 neutralizing antibodies.

**Conclusions and Relevance:** This study demonstrates that humoral responses to SARS-CoV-2 infection are robust on longer time-scales, including those arising from milder infections.

However, the magnitude and durability of the antibody response after natural infection was lower and more variable in younger participants who did not require hospitalization for COVID-19. These findings support vaccination against SARS-CoV-2 in all suitable populations including those individuals that have recovered from natural infection.

## INTRODUCTION

The immune correlates of protection against severe acute respiratory syndrome coronavirus 2 (SARS-CoV-2) infection and coronavirus disease 2019 (COVID-19) are unknown. However, the development of detectable humoral immunity is likely a predictive surrogate of protection^1,2^. The presence of broadly neutralizing serum antibodies five to eight months after SARS-CoV-2 infection have been documented by several groups^3-10^. Cases of symptomatic COVID-19 following re-infection with SARS-CoV-2 have been reported but are infrequent^11-15^, and recent studies have highlighted a correlation between the presence of SARS-CoV-2 antibodies and decreased risk of reinfections^16,17^.

The magnitude of the antibody response to SARS-CoV-2 infection has been positively correlated with COVID-19 severity^18-25^, but the confounding effect of age on this association remains unresolved^26-28^. Even less understood is whether cross-reactive seasonal human coronavirus (HCoV) antibodies correlate with the kinetics of SARS-CoV-2 humoral responses across acute and post-acute timescales after SARS-CoV-2 infection^29-32^. Pre-existing HCoV antibodies that cross-react with but do not cross-neutralize SARS-CoV-2 have been detected^30,33-36^, and recent infection with HCoVs has been correlated with reduced COVID-19 severity^37^.

Here, we demonstrate the persistence of SARS-CoV-2 IgG binding and neutralizing responses out to twelve months in participants enrolled in a prospective study at seven military treatment facilities (MTFs) across the U.S. from March 2020 to March 2021. MTFs provide care for active duty servicemembers, dependents and military retirees, including a range of ages and demographics that is broadly representative of the civilian U.S. population. Study participants were followed for up to twelve months allowing analyses to identify correlates of long humoral immune durability to SARS-CoV-2. The aims are to (i) describe the magnitude and durability of SARS-CoV-2 antibody response for one year after natural infection, and (ii) identify correlates of SARS-CoV-2 antibody response, including COVID-19 severity, age, and antibody profiles to HCoVs.

## METHODS

### Study population, setting, participant enrollment and sera collection

Participants were enrolled and serum samples were collected in the Epidemiology, Immunology, and Clinical Characteristics of Emerging Infectious Diseases with Pandemic Potential (EPICC) protocol: a prospective, longitudinal study of COVID-19. The protocol was approved by the Uniformed Services University Institutional Review Board (IDCRP-085), and all subjects or their legally authorized representative provided informed consent to participate. Participants were enrolled at seven MTFs across the United States, including Walter Reed National Military Medical Center (Bethesda, MD), Brooke Army Medical Center (San Antonio, TX), Naval Medical Center San Diego (San Diego, CA), Naval Medical Center Portsmouth (Portsmouth, VA), Madigan Army Medical Center (Tacoma, WA), Fort Belvoir Community Hospital (Fort Belvoir, VA) and Tripler Army Medical Center (Honolulu, HI). Eligible participants included individuals with laboratory-confirmed SARS-CoV-2 infection by nucleic acid amplification test (NAAT), individuals with SARS-CoV-2-like illness, and individuals who were tested following a high risk exposure to a SARS-CoV-2 positive person or screening surrounding travel. Blood specimens were collected at enrollment, and then at seven, fourteen, and twenty-eight days, and subsequently at three, six and twelve months after enrollment.

Antibody results from SARS-CoV-2 PCR-positive (n=505) and SARS-CoV-2 PCR-negative (n=92) participants were included in the evaluation of humoral response to SARS-CoV-2 infection. From these participants, we analyzed spike protein IgG binding in a serial collection of 764 serum samples from 250 (outpatients n= 192, inpatients n= 58) participants who were followed through six and twelve months-post-enrollment. Six months serum samples from these participants were collected at a median 188 days post-symptom onset (dpso), IQR= 15. Of these 250 participants, 19 (outpatients, n= 11; inpatients= 8) had available sera drawn twelve months from the onset of symptoms and prior to vaccination, allowing long-term monitoring of IgG duration (eFigure 1). Serum samples collected from individuals after the administration of COVID-19 vaccinations were excluded from this analysis of antibody responses to natural infection. To characterize the durability of the neutralizing antibody response to SARS-CoV-2, paired sera from 72 participants who had serum samples collected during early convalescence (median 36 dpso, IQR= 14.50) and at six months-post symptom onset collected from September to October 2020 were evaluated by a SARS-CoV-2 S-pseudovirus neutralization test (SNT) and an authentic wild-type SARS-CoV-2 virus neutralization test (VNT). Twelve months-post sera collected in March 2021 from 7 inpatients and 4 outpatients were further evaluated by SNT.

**Figure 1.**
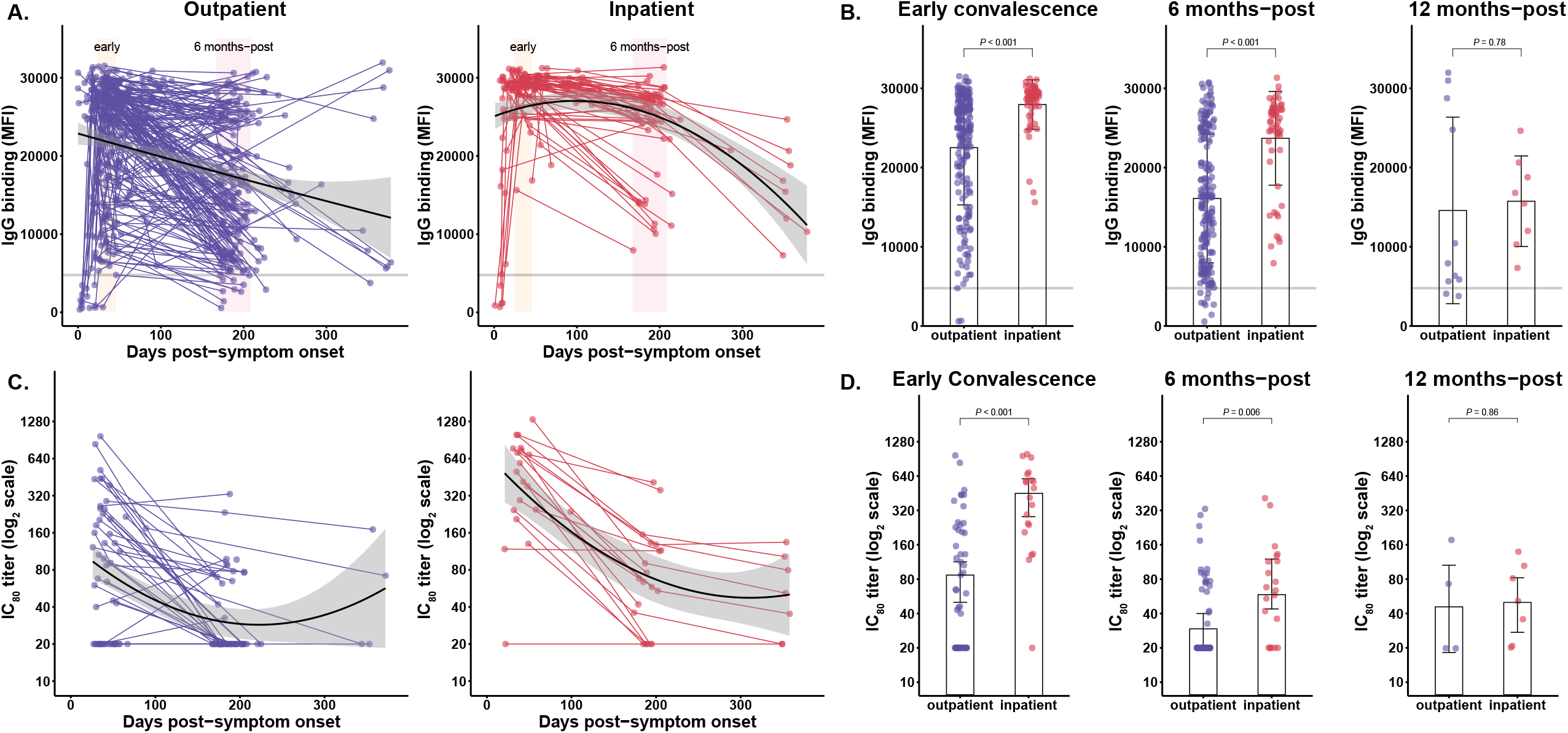
Evaluation of the magnitude and duration of the antibody response and COVID-19 clinical phenotype. Non-linear regressions were used to compare IgG responses from **(A)** outpatients (n=192) and inpatients (n=58). Longitudinal samples for subjects are connected by lines; second order polynomial curves were fit to inpatient (red) and outpatient (blue) groups; 95% CIs are shaded gray. A horizontal line indicates the indeterminate range between SARS-CoV-2 positive (>4774) and negative (<4144) IgG; MFI, median fluorescence intensity. Two distinct shaded regions highlighted early convalescence (yellow) and 6 months-post (pink) windows. **(B)** Early convalescence (median 35 dpso), six months-post (median 188 dpso) and twelve months-post (median 357 dpso) IgG responses were compared between outpatients and inpatients; error bars indicate the geometric mean and 95% CI. **(C)** Longitudinal SNT neutralizing antibody responses of outpatients (n=54) and inpatients (n=20). (**D)** Early convalescence and six months-post SNT neutralizing antibodies were compared by hospitalization status. *P-*values were determined by unpaired t-test with Welch’s correction, α= 0.05; error bars indicate the geometric mean and 95% CI.

### Multiplex microsphere-based immunoassay screening procedures

Detailed experimental procedures of SARS-CoV-2 and HCoV spike protein-based multiplex microsphere immunoassays have been previously described^38-40^ and are described further in the Supplementary Appendix (eMethods). Briefly, diluted serum and capillary blood samples were tested in technical duplicates. Antigen-antibody complexes were analyzed on Bio-Plex 200 multiplexing systems (Bio-Rad, Hercules, CA) for IgG binding and median fluorescence intensity (MFI) values are reported.

### SARS-CoV-2 S-pseudovirus production and neutralization (SNT)

The spike (S) sequence from SARS-CoV-2 isolate Wuhan-Hu-1 (GenBank accession: YP_009724390.1) was used to construct lentiviral pseudoviruses for the neutralization assays, as described previously^41^. Additional details are provided in the (eMethods), briefly, pseudovirus titers were measured by infecting 293T-ACE2.TMPRSS2 cells. Pseudovirus titers were determined as relative luminescence units per milliliter of pseudovirus supernatants (RLU/ml). The antibody dilution causing a 50% and 80% reduction (inhibitory concentration, IC) of vector-expressed luciferase compared to control (IC_50_- and IC_80_-neutralizing antibody titer, respectively) was calculated with nonlinear regression using GraphPad Prism. Data reported were averages from at least two independent experiments.

### Wild-type SARS-CoV-2 plaque reduction neutralization tests (VNT)

VNT antibody titers were determined by plaque reduction neutralization test (PRNT) as previously described with modifications^42^. Details of experimental procedures are included in the Supplementary Appendix (eMethods). SARS-CoV-2 (USA WA1/2020, BEI Resources cat # NR-52281) and serum samples were incubated for one hour then incubated with Vero-81 cells (ATCC cat NoCRL-1587). Cutoffs for 80% PRNT titers (PRNT_80_) were determined on each plate. Wells with an OD_405_ less than 20% of the mean value of nine virus only controls, plus one standard deviation, were considered neutralizing.

### Statistical analysis of humoral response correlates

Log-scale transformations were applied to all SARS-CoV-2 IgG binding and neutralization antibody datasets to explore normality and parametric or non-parametric were applied as indicated. For VNT PRNT80 titers, zero values were changed to 0.01 prior to log10-transformation and nonparametric unpaired Mann-Whitney tests were performed. Generally, second order polynomial curves were the preferred non-linear regression model (α= 0.05) and these best-fit curves with confidence intervals are shown in all graphs. Exponential phase-decay analyses were used to explore antibody half-life (*T*_1/2_) trends utilizing subjects with ≥2 longitudinal sera samples, and, based on best-fit, either a one-phase or two-phase decay model was preferred. When single models for all the datasets were not preferred or a best-fit single curve was ambiguous, a robust fit without curve fitting was applied and the mean of all subjects’ individual *T*_1/2_ was calculated; in several instances *T*_1/2_ exceeded 1000 days and were reported as >1000. We used Brown-Forsythe and Welch’s ANOVA to compare age-stratified log10-transformed IgG binding MFI data and adjusted for multiple comparisons through use of the Dunnet’s multiple T3 comparison test. Box-Cox transformations were applied to HCoV IgG binding MFI values to normalize the data and parametric t-tests were performed. Multivariate linear regression models were used to compare MFI among age groups and by hospitalization status (with interaction term), and separate models were run for samples collected in the early convalescence period and at six months-post. Figures were generated and statistical analyses were performed in GraphPad Prism version 9.0.2 and RStudio version 4.0.2 software (R Foundation for Statistical Computing)^43^.

## RESULTS

### Demographic and hospitalization status of EPICC participants

Over half of the participants were 18-44 years of age or male. The racial distribution of participants was non-Hispanic white (44.3%), followed by Hispanic (31.2%) and African-American (14.1%) (Table 1). Participants were classified according to the maximum severity reported during follow-up as hospitalized (inpatients) or outpatients. Participants were stratified into three age groups: 18-44, >44-64 and ≥65 years old. The median age of inpatient and outpatient participants was 58.2 (interquartile range (IQR)= 16.3 years) and 43.3 (IQR= 24.4) years, respectively.

**Table 1.**
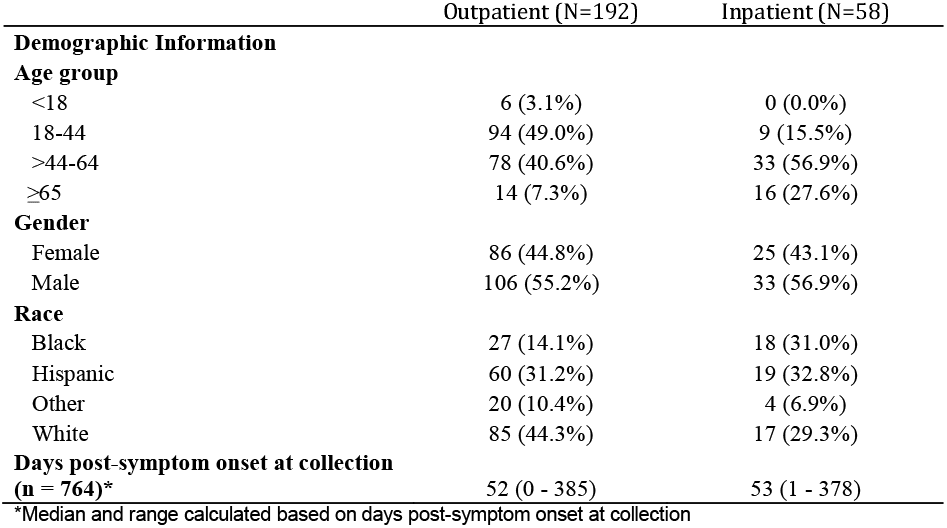
Baseline characteristics of participants included in the longitudinal study of antibody responses.

### SARS-CoV-2 binding and neutralizing antibody responses differ by COVID-19 severity

We observed 95% (183/192) of outpatients and 100% of inpatients (58/58) remained seropositive at six months-post, and 9/11 outpatients and 8/8 inpatients remained seropositive at 12 months-post symptom onset. A one-phase decay of the IgG response of inpatients calculated a *T*_1/2_ >1000 days (Figure 1A). IgG responses displayed greater heterogeneity among outpatients than inpatients and a one-phase decay curve modeled a *T*_1/2_= 1232 days (Figure 1A). Next, we sought to investigate whether the magnitude or duration of the IgG response was associated with COVID-19 clinical disease severity as determined by hospitalization status. For this analysis, magnitude was explored as IgG responses recorded during early convalescence for each participant across all longitudinal sera collections and the durability of the antibody response was assessed with sera collected six and twelve months-post symptom onset. Geometric mean IgG levels during early convalescence and six months-post-infection were significantly higher in inpatients than in outpatients (early convalescence: inpatients= 27,646 MFI [95% Confidence Interval (CI): 26,688-28,639], outpatients= 20,587 MFI [CI:19,057-22,241], *P <*0.001; six months-post-infection: inpatients= 22,694 MFI [95% CI: 19,967-25,792], outpatients= 13,559 MFI [95% CI: 12,343-14,895], *P* <0.001) (Figure 1B). By twelve months-post we found no differences in geometric mean IgG binding between inpatients (14,755 [95% CI: 11,181-19,472]) and outpatients (10,588 [95% CI: 6,421-17,460]) (*P=* 0.78). In addition to MFI as a measurement of IgG binding, we determined anti-SARS-CoV-2 IgG endpoint titers. Again, we found that the geometric mean of endpoint titers (GMT) were significantly higher for inpatients than outpatients during early convalescence (inpatients= 13,029 [95% CI: 9375-18,108], outpatients= 3240 [95% CI: 2323-4518]) (eFigure 2A), and six months-post (inpatients= 8268 [95% CI: 5323-12,843], outpatients= 2216 [95% CI: 1654-2970]) (eFigure 2B).

**Figure 2.**
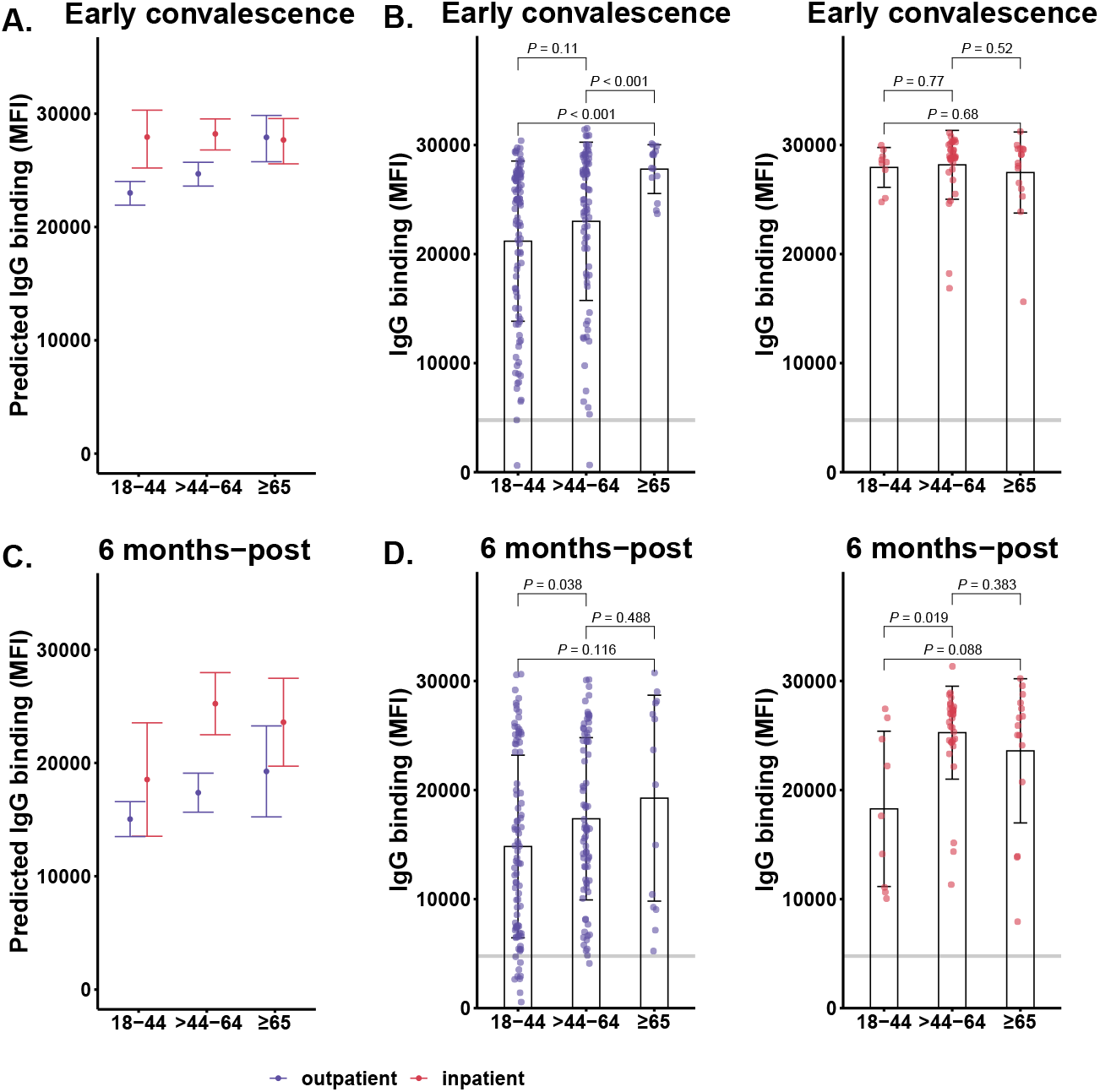
The magnitude and durability of IgG-binding responses are associated with COVID-19 severity and age. **(A)** Multivariate linear regression analysis of outpatient and inpatient IgG responses and **(B)** hospitalization status stratified by age groups, outpatients, 18-44 (n=94), >44-64 (n=78), ≥65 (n=14), and inpatients, 18-44 (n=9), >44-64 (n=33), ≥65 (n=16) during early convalescence. A horizontal line indicates cutoff for positive/negative IgG; MFI, median fluorescence intensity. Statistical significance were determined by unpaired t-test with Welch’s correction, α = 0.05; error bars indicate the geometric mean and 95% CI. **(C-D)** Six months-post IgG responses were compared between age-stratified outpatients and inpatients.

Next, sera were assessed for neutralizing antibodies by SNT; IC_80_ titers are shown in Figures 1 and 2, while IC_50_ titers are provided in eFigure 3A-C. A one-phase decay modeled inpatient *T*_1/2_ neutralizing antibody responses of 88 days and a two-phase decay of outpatient neutralizing antibody responses calculated a mean fast/slow-*T*_1/2_ of 77/132 days (Figure 1C). The neutralizing antibody GMT was greater for inpatients than outpatients during both early convalescence, 378 [95% CI: 246-580] versus 83 [95% CI: 59-116] (P <0.001), and six months-post, 65 [95% CI: 43-98] versus 33 [95% CI: 26-40] (P= 0.006), although these differences were not observed by twelve months-post (Figure 1D). These significant associations between COVID-19 severity, and IC_80_ neutralizing antibody kinetics and durability were also observed with IC_50_ titers (eFigure 3A-C).

**Figure 3.**
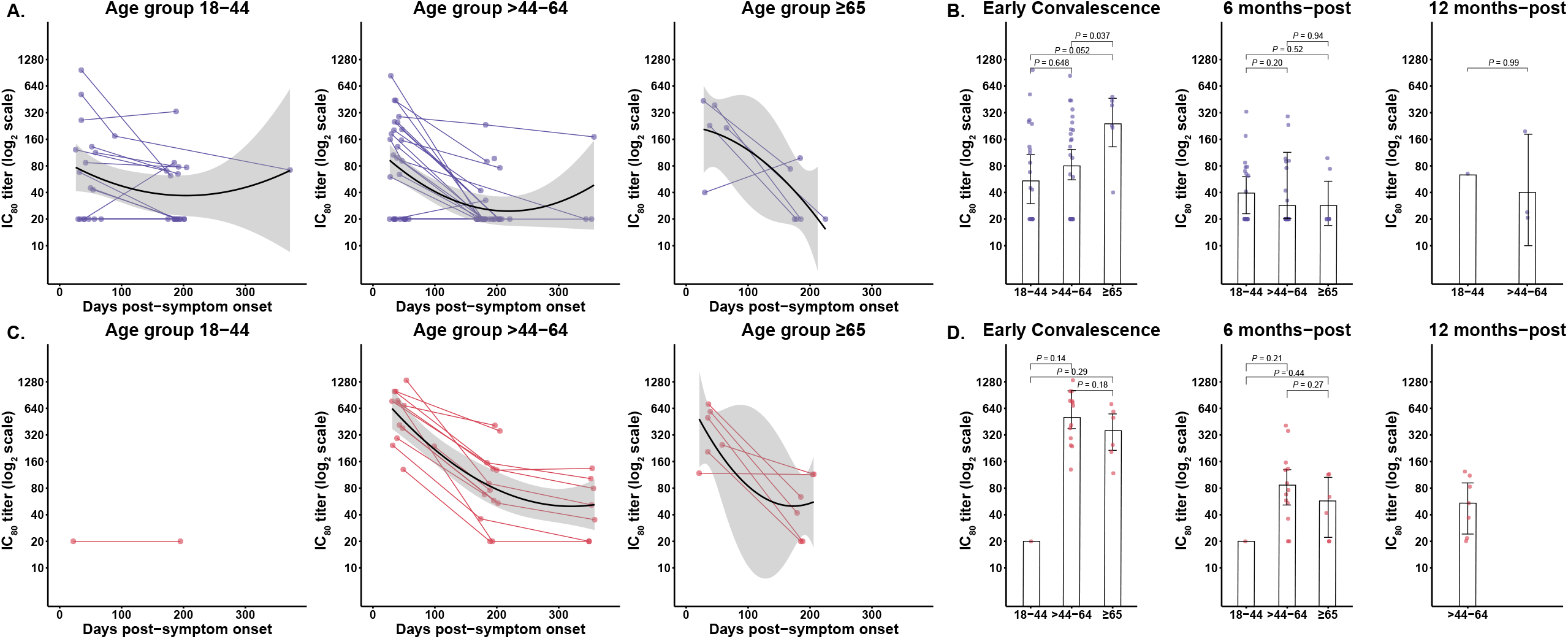
The magnitude and durability of neutralizing antibody responses are associated with COVID-19 severity and age. **(A)** Longitudinal SNT measurement of neutralizing antibodies in outpatient age groups, 18-44 (n=18), >44-64 (n=29) and ≥65 (n=6); longitudinal samples are connected by lines, second order polynomial curves and 95% CIs are shaded gray. **(B)** Early convalescence and six months-post SNT measured neutralizing antibodies compared between outpatient age groups. **(C)** Longitudinal SNT measurement of neutralizing antibodies in outpatient age groups, 18-44 (n=1), >44-64 (n=13) and ≥65 (n=6). **(D)** Early convalescence and six months-post SNT measured neutralizing antibodies compared between inpatient age groups. Statistical significance were determined by unpaired t-test with Welch’s correction, α = 0.05; error bars indicate the geometric mean and 95% CI.

Recapitulating the durability, magnitude, and correlates of humoral immune response to SARS-CoV-2 across different populations with different neutralization assays remains a critical goal^44^. Antibody neutralization was further characterized by a wild-type SARS-CoV-2 VNT. Endpoint titers in VNT correlated significantly and had a modest coefficient strength with SNT titers (Spearman’s ρ= 0.77, *P <*0.001) (eFigure 4A). A one-phase decay of VNT neutralizing antibody responses calculated a *T*_1/2_ of 106 and 29 days for inpatients and outpatients, respectively (eFigure 4B-C). The magnitude and durability of VNT GMT was also different between inpatients and outpatients during early convalescence (inpatients=52 [95% CI: 14-198], outpatients=11 [95% CI: 4-29], *P* <0.001) and six months-post (inpatients=14 [95% CI: 3-71], outpatients=2 [95% CI: 0.5-5], *P*= 0.02) (eFigure 4D).

**Figure 4.**
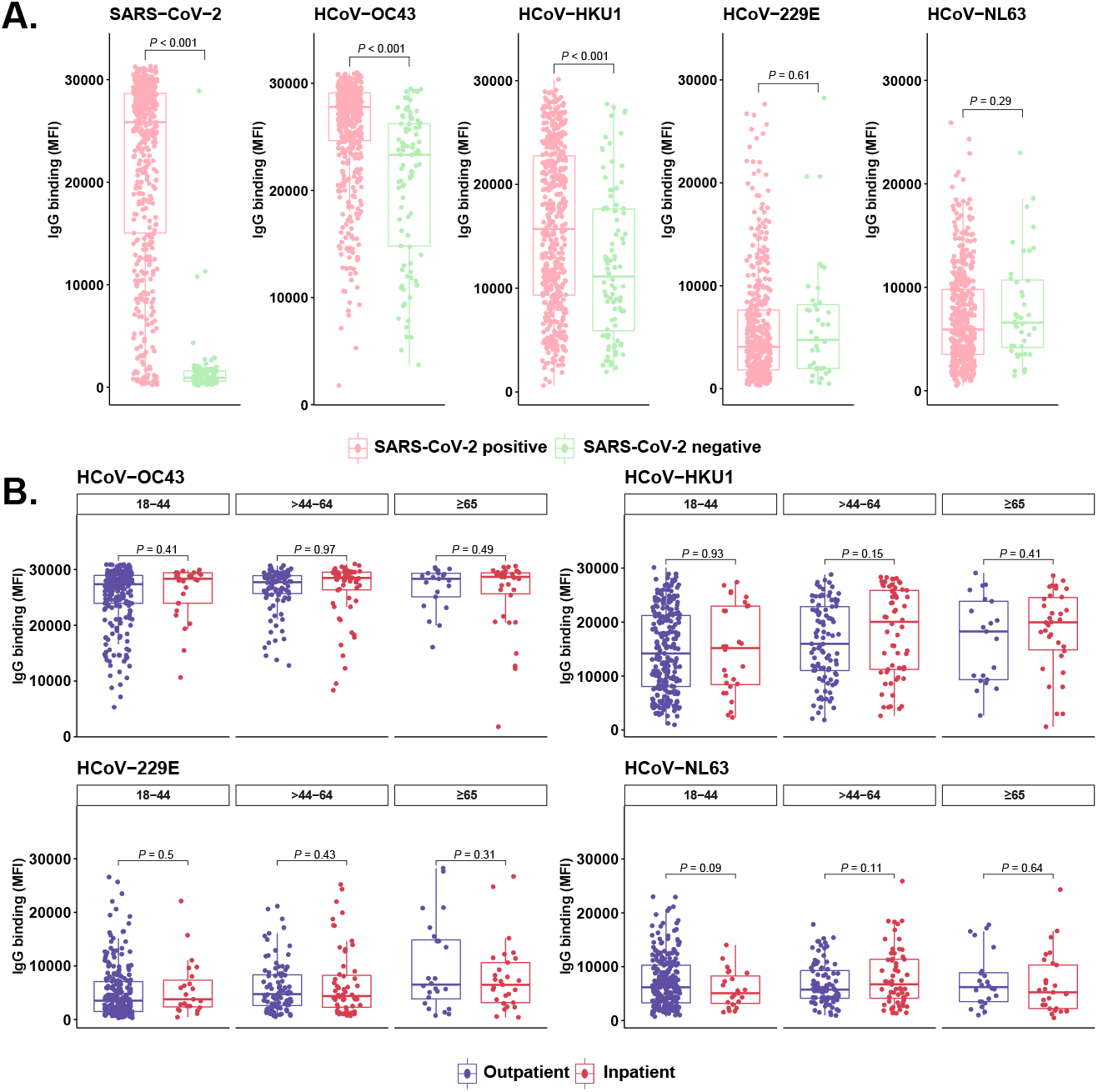
Seasonal HCoV antibody responses are not associated with COVID-19 clinical outcomes. **(A)** IgG binding levels of SARS-CoV-2 and seasonal HCoV-OC43, HCoV-HKU1, HCoV-229E, HCoV-NL63 detected in SARS-CoV-2 PCR-positive (n=505) and SARS-CoV-2 PCR-negative (n=92) samples. **(B)** Stratified SARS-CoV-2 positive samples (n=505) into age groups (18-44, >44-64, and ≥65 years old) and COVID-19 severity (outpatient vs. inpatient). MFI, median fluorescence intensity; dpso is from zero to twelve months; boxplots denote median, first quartile (25^th^ percentile) and third quartile (75^th^ percentile); statistical significance was determined by unpaired t-test with Welch’s correction, α = 0.05.

### Age correlation with antibody durability may be explained by age-specific clinical severity

Because age is associated with hospitalization status, we used a multivariate regression model to explore antibody magnitude and durability on the basis of COVID-19 severity and age (groups: 18-44, >44-64, and ≥65-years-old). This analysis revealed that during early convalescence IgG levels were higher for all inpatient participants, and increased with age for outpatients with significantly higher IgG-binding levels in ≥65-years-old outpatients that was comparable to ≥65-years-old inpatients (Figure 2A). Furthermore, significant differences in IgG-binding levels were noted between outpatients in 18-44-years-old (19,124 MFI [95% CI: 17,058-21,439, *P* <0.001) and >44-64-years-old groups (20,897 MFI [95% CI: 18,404-23,728], *P <*0.001) compared to the ≥65-years-old group (27,703 MFI [95% CI: 26,401-29,069]) (Figure 2B). By six months-post, IgG levels remained higher for inpatients across age groups than the outpatients (Figure 2C), and significantly so for the >44-64-years-old (24,789 MFI [95% CI: 22,947-26,779], *P=* 0.019) compared to the 18-44 years-old age group (Figure 2D). Additionally, no differences in the IgG response were detected by twelve months-post (eFigure 5A). The IgG *T*_1/2_ of outpatient age groups 18-44-year-old, >44-64-year-old and ≥65-year-old, were >1000, 230, and 143 days, respectively (eFigure 5B). Compared to age-grouped outpatients, IgG *T*_1/2_ of inpatient age groups were slower, >1000 days for all 18-44-year-old, >44-64-year-old and ≥65-year-old age groups (eFigure 5C).

Next, we compared age-stratified neutralizing antibody titers across outpatients and inpatients. In outpatient 18-44, >44-64 and ≥65 age-groups, neutralizing antibodies one-phase decay *T*_1/2_ were 16, 34, and 21 days, respectively (Figure 3A). Strikingly, we noted a higher magnitude during early convalescence in outpatients ≥65-years-old (GMT: 233 [95% CI: 111-489]) compared to 18-44 (GMT: 67 [95% CI: 37-120], *P=* 0.052) and >44-64 (GMT: 80 [95% CI: 50-127], (*P*= 0.037) years-old groups (Figure 3B). However, this difference was not observed by six months-post, correlating with the short *T*_1/2_ in the ≥65-years-old group (Figure 3B). The slowest one-phase decay *T*_1/2_ was observed in the inpatient ≥65-years-old group, 84 days (Figure 3C), and when comparing inpatient neutralizing antibodies during early convalescence, higher GMT were observed in the >44-64 and ≥65-years-old groups, 505 [95% CI: 346-738] (*P=* 0.14) and 328 [95% CI: 187-576] (*P=* 0.18), respectively (Figure 3D). These results appear to suggest that the correlation between age and early humoral response is confounded by age-specific severity of SARS-CoV-2 infection, consistent with other findings^45^.

### Seasonal HCoV antibody responses are not correlated with COVID-19 outcomes or the development of neutralizing antibodies

We first explored whether subjects with PCR-confirmed SARS-CoV-2 infection possessed higher levels of HCoV spike protein reactive antibodies as compared to SARS-CoV-2 negative subjects. Higher levels of HCoV-OC43 and HCoV-HKU1 reactive IgG, but not of HCoV-229E and HCoV-NL63 were observed in SARS-CoV-2-positive subjects during early convalescence across age groups with mild to severe COVID-19 (Figure 4A). Further, we identified a positive correlation and distinct clustering of maximum IgG levels between SARS-CoV-2 and seasonal betacoronaviruses (HCoV-OC43 and HCoV-HKU1) that was related to dpso (eFigure 6A-B), but only very weak relationships with the seasonal alphacoronaviruses (HCoV-229E and HCoV-NL63) (eFigure 6C-D). To examine the clinical correlation between HCoV antibody responses and COVID-19 severity, subjects were again stratified by age and clinical phenotype; we observed no significant correlation with HCoV peak antibody responses (Figure 4B). Finally, we sought to determine whether the induction of cross-reactive HCoV antibodies following SARS-CoV-2 infection were associated with the magnitude or durability of neutralizing antibodies to SARS-CoV-2. The magnitude of HCoV-OC43 and HCoV-HKU1 IgG titers during early convalescence was not significantly associated with SARS-CoV-2 neutralizing antibody responses during either early convalescence or six months-post symptom onset (eFigure 7A-D).

## DISCUSSION

In this study, we have demonstrated that SARS-CoV-2 binding IgG and neutralizing antibodies remained detectable for up to one year in subjects following mild and moderate to severe COVID-19. Further, we corroborated that the magnitude and durability of humoral immune response are positively correlated, reflected by both *T*_1/2_ and levels of binding IgG and neutralizing antibody detected at time periods during early convalescence and six months-post symptom onset ^46,47^. This may be due to robust stimulation of humoral immunity with failure to control infection via innate responses.

Notably, when we controlled for hospitalization status, older age was positively correlated with robust positive antibodies and neutralizing antibody responses. This suggests a lack of immunosenescence driving waning humoral responses or seroreversion as all instances of seroreversion between six to twelve months-post symptom onset occurred in outpatient participants <65 years old (median age 30, Q1=26, Q3=43). Although, the association between age and disease severity may confound this observation. The interaction between age, severity and adaptive responses is complex^48,49^; we noted that age ≥65 years was significantly associated with the magnitude and durability of IgG responses for outpatients, whereas no differences were found for inpatients across the age groups. However, sample size was smaller in the inpatient group so this observation needs to be investigated further. Additionally, the magnitude of the early neutralizing antibody response increased incrementally in outpatients and inpatients age groups >44 years old. Interestingly, no significant differences in neutralizing antibody levels were observed across age groups by six months after symptom onset.

When we assessed HCoV seroresponses in our cohort, we found no association with the presence of antibodies to seasonal HCoVs and COVID-19 severity or with the development of SARS-CoV-2 neutralizing antibodies. The induction of antibodies cross-reactive with HCoV spike proteins after SARS-CoV-2 infection and boosted HCoV-HKU1 and HCoV-OC43 responses were observed, implying that highly conserved betacoronavirus spike protein epitopes, possibly conformation-dependent, are cross-reactive^50^. This conclusion is supported by prior observations that conserved regions of the SARS-CoV-2 spike protein S2 subunit have been shown to stimulate specific memory B cell repertoires^51,52^. Although this investigation is limited by the lack of baseline pre-SARS-CoV-2 infection sera, we also showed that boosted HCoV-OC43 and HCoV-HKU1 memory responses were not associated with COVID-19 clinical outcomes nor detrimental to the *de novo* development of SARS-CoV-2 neutralizing antibodies^30^.

Our finding of variable waning yet persistent neutralization titers across participants groups is consistent with other longitudinal studies^7,53-55^, however neutralization presents only one facet of long term SARS-CoV-2 immunity. Memory B cells specific to the SARS-CoV-2 spike receptor-binding domain, which are immunodominant and responsible for 90% of neutralizing activity^56^, have been detected even when circulating serum neutralizing antibodies have waned below detectable limits^7,55^.

Our results add to the growing body of literature that suggests humoral immunity following natural infection with SARS-CoV-2 is long lived, including out to one year post-infection. However, the magnitude and durability of SARS-CoV-2 antibody response was lower and more variable in younger participants (<65 years old) who experienced less severe COVID-19 and did not require hospitalization. These findings suggest that implementation of vaccination against SARS-CoV-2 infection in all suitable populations, including those individuals that have recovered from natural infection, would be prudent because vaccine induced immunity to SARS-CoV-2 will likely be more long-lived than that elicited from mild COVID-19. Additional studies will also be critical to further examine the protective role and durability of antibody responses following SARS-CoV-2 re-infection and/or vaccination up to and beyond one year.

## Data Availability

The data that support the findings of this study are available from the corresponding author upon reasonable request.

## DECLARATIONS

This research protocol, IDCRP-085, was approved by the Uniformed Services University Institutional Review Board.

## STATEMENT OF ETHICS

The referenced human subjects protocol (IDCRP-085) was approved by the Uniformed Services University Institutional Review Board and participating sites. All subjects or their legally authorized representative provide written or verbal informed consent using approved documents and procedures; the consent forms include clauses allowing use of specimens for investigations including those conducted in this study.

## DISCLAIMER

The contents of this publication are the sole responsibility of the author(s) and do not necessarily reflect the views, opinions, or policies of the Uniformed Services University (USU), the Henry M. Jackson Foundation for the Advancement of Military Medicine, Inc. (HJF), National Institutes of Health or the Department of Health and Human Services, Brooke Army Medical Center, Naval Medical Center Portsmouth, Tripler Army Medical Center, Naval Medical Center San Diego, Madigan Army Medical Center, Walter Reed National Military Medical Center, the U.S. Army Medical Department, the U.S. Army Office of the Surgeon General, the US Department of Defense (DoD), the Departments of the Air Force, Army or Navy, or the U.S. Government. Mention of trade names, commercial products, or organization does not imply endorsement by the U.S. Government. The investigators have adhered to the policies for protection of human subjects as prescribed in 45 CFR 46. A number of the co-authors are employees of the U.S. Government or military service members. This work was prepared as part of their official duties. Title 17 U.S.C. §105 provides that ‘Copyright protection under this title is not available for any work of the United States Government.’ Title 17 U.S.C. §101 defines a U.S. Government work as a work prepared by a military service member or employee of the U.S. Government as part of that person’s official duties.

## ACKNOWLEDGEMENTS

We thank Dominic Esposito, Matthew Drew, Jennifer Melhako, Kelly Snead, Vanessa Wall, John-Paul Denson, Simon Messing, and William Gillette (Protein Expression Lab, FNCLR) for provision of HCoV-HKU1 and HCoV-OC43 spike proteins and excellent technical assistance. Additionally, we thank Shreshta Phogat (USU, HJF) for technical assistance. We also thank the members of the EPICC COVID-19 Cohort Study Group for their many contributions in conducting the study and ensuring effective protocol operations. The following members were all closely involved with the design, implementation, and oversight of the study.

*Brooke Army Medical Center, Fort Sam Houston, TX:* J. Cowden; D. Lindholm; A. Markelz; K. Mende; T. Merritt; R. Walter; CPT T. Wellington

*Carl R. Darnall Army Medical Center, Fort Hood, TX:* MAJ S. Bazan

*Fort Belvoir Community Hospital, Fort Belvoir, VA:* L. Brandon; N. Dimascio-Johnson; MAJ E. Ewers; LCDR K. Gallagher; LCDR D. Larson; MAJ M. Odom

*Henry M. Jackson Foundation, Inc*., *Bethesda, MD:* P. Blair; D. Clark

*Madigan Army Medical Center, Joint Base Lewis McChord, WA:* LTC C. Colombo; R. Colombo; CPT C. Conlon; CPT K. Everson; LTC P. Faestel; COL T. Ferguson; MAJ L. Gordon; LTC S. Grogan; CPT S. Lis; COL C. Mount; LTC D. Musfeldt; MAJ R. Sainato; C. Schofield; COL C. Skinner; M. Stein; MAJ M. Switzer; MAJ M. Timlin; MAJ S. Wood

*Naval Medical Center Portsmouth, Portsmouth, VA:* G. Atwood; R. Carpenter; LCDR C. Eickhoff; CAPT K. Kronmann; T. Lalani; LCDR T. Lee; T. Warkentien

*Naval Medical Center San Diego, San Diego, CA:* CAPT J. Arnold; CDR C. Berjohn; S. Cammarata; LCDR S. Husain; LCDR A. Lane; CAPT R. Maves; J. Parrish; G. Utz

*Tripler Army Medical Center, Honolulu, HI:* S. Chi; MAJ E. Filan; K. Fong; CPT T. Horseman; MAJ M. Jones; COL A. Kanis; LTC A. Kayatani; MAJ W. Londeree; LTC C. Madar; MAJ J. Masel; MAJ M. McMahon; G. Murphy; COL V. Nguay; MAJ E. Schoenman; C. Uyehara; LTC R. Villacorta Lyew

*Uniformed Services University of the Health Sciences, Bethesda, MD:* B. Agan; C. Broder; CAPT T. Burgess; COL K. Chung; C. Coles; C. English; M. Haigney; COL P. Hickey; E. Laing; LTC J. Livezey; A. Malloy; COL T. Oliver; E. Parmelee; S. Pollett; S. Richard; J. Rozman; M. Sanchez; A. Scher; CDR M. Simons; A. Snow; D. Tribble

*United States Air Force School of Medicine, Dayton, OH:* A. Fries

*Walter Reed National Military Medical Center, Bethesda, MD:* A. Ganesan; D. Gunasekera; MAJ N. Huprikar; M. Oyeneyin

*William Beaumont Army Medical Center, El Paso, TX:* CPT M. Banda; CPT B. Davis; MAJ T. Hunter; CPT O. Ikpekpe-Magege; CPT S. Kemp; R. Mody; R. Resendez; COL M. Wiggins

*Womack Army Medical Center, Fort Bragg, NC:* B. Barton; D. Hostler; C. Maldonado

