## Supplementary Appendix for "SARS-CoV-2 antibodies remain detectable 12 months after infection and antibody magnitude is associated with age and COVID-19 severity"

Laing *et al.*

1. **eMethods**
2. **eFigures**
    a. **eFigure 1**
    b. **eFigure 2**
    c. **eFigure 3**
    d. **eFigure 4**
    e. **eFigure 5**
    f. **eFigure 6**
    g. **eFigure 7**

This supplemental material has been provided by the authors to give readers additional information about their work.

### 1. eMethods

**Multiplex microsphere-based immunoassay screening procedures**

Serum samples were collected from venipuncture in serum separator tubes, processed and stored at -80 °C in 250 µL aliquots until use. As previously validated, neat human serum samples (1.25 µL) were diluted 1:400 in 1X PBS and dried blood spots collected by at-home capillary blood (Mitra® microsampling device, Neoteryx, Torrance, CA) were eluted 1:20 then diluted 1:10 and treated similarly to serum samples. All blood specimen samples were heat inactivated at 60 ˚C for 30 min after dilutions and tested on technical duplicate plates. Blood specimens were incubated with a master mix of SARS-CoV-2, HCoV-OC43, HCoV-HKU1, HCoV-229E, and HCoV-NL63 native-like spike protein ectodomain trimers coupled magnetic microspheres (BioRad, Hercules, CA). After a 45 minute incubation with agitation (900 rpm), plates were washed with PBS-Tween20 (0.05%) and 100 µL of biotinylated cross-absorbed anti-human IgG (Thermo Fisher Scientific, Waltham, MA) diluted in 1X PBS-T (1:5000) was added to each well, and plates were incubated for 45 minutes with agitation. Lastly, after washing, streptavidin-phycoerythrin was diluted 1:1000 in PBS-T, and 100 µL were added to each well and plates were incubated for 45 min with agitation (900 rpm). Plates were washed, and microspheres were resuspended with 100 µL PBS-T per well then analyzed on Bio-Plex 200 multiplexing systems (Bio-Rad) and median fluorescence intensity (MFI) values for samples are reported as the PBS adjusted average from duplicate plates. Antibody testing was blind to descriptive data, including demographic data, SARS-CoV-2 PCR status, clinical phenotype and enrollment/collection dates. IgG binding MFI values <4144 were deemed negative and those >4774 were deemed positive; MFI values between the 4144 to 4774 range are indeterminate. SARS-CoV-2 IgG sensitivity and specificity of this assay 7-28 days post-symptom onset (dpso) are 94.44% and 100%, respectively.

**SARS-CoV-2 S-pseudovirus production and neutralization (SNT)**

The spike (S) sequence from SARS-CoV-2 isolate Wuhan-Hu-1 (GenBank accession: YP_009724390.1) was used to construct lentiviral pseudoviruses for the neutralization assays, as described previously for pseudoviruses bearing other viral glycoproteins [1]. Briefly, 5µg of pCMVΔR8.2, 5µg of pHR'CMVLuc and 0.5µg of S expression plasmids were co-transfected in 293T cells. Pseudovirus supernatants were collected approximately 48 hours post-transfection, filtered through a 0.45 µm low protein binding filter, and used immediately or stored at -80ºC. Pseudovirus titers were measured by infecting 293T-ACE2.TMPRSS2 cells, which stably express human angiotensin converting enzyme 2 (ACE2) and transmembrane serine protease 2 (TMPRSS2), for 48 hours prior to measuring luciferase activity (luciferase assay reagent, Promega, Madison, WI). Pseudovirus titers were determined as relative luminescence units per milliliter of pseudovirus supernatants (RLU/ml). The antibody dilution causing a 50% and 80% reduction (inhibitory concentration, IC) of vector-expressed luciferase compared to control ($IC_{50}$- and $IC_{80}$-neutralizing antibody titer, respectively) was calculated with nonlinear regression using GraphPad Prism. Data reported were averages from at least two independent experiments.

**Wild-type SARS-CoV-2 plaque reduction neutralization tests (VNT)**

Patient serum was heat inactivated at 56°C for 30 minutes and serially diluted into complete MEM. 100 µl of the diluted serum was transferred, in triplicate, to a 96-well tissue culture plate containing 50 µl of MEM with 200 $TCID_{50}$ of SARS-CoV-2 (USA WA1/2020, BEI Resources cat # NR-52281). SARS-CoV-2 (USA WA1/2020, BEI Resources cat # NR-52281) and serum samples were incubated at 37°C with 5% CO 2 for one hour, after which, 100 µl of media containing 1×10 5 /ml of Vero-81 cells (ATCC cat NoCRL-1587) was added to each well and the plates were incubated at 37°C with 5% CO2 for 72 hours. As controls, nine wells contained Vero cells and virus, and three wells contained Vero cells only. Following incubation, plates were fixed with the addition of 200 µl ice cold methanol/ethanol (50% vol/vol) to each well and incubated at room temperature for 1 hour. After fixation, an ELISA targeting the SARS-CoV-2 spike protein was conducted. Here, wells were blocked for one hour in blocking buffer (1X PBS plus 0.1% TWEEN-20 with 5% milk) and washed 3X. SARS-CoV-2 spike protein antigen-antibody complexes were detected by ELISA of SARS-CoV-2 spike protein polyclonal rabbit sera (1:500)(produced in house),goat anti-rabbit IgG (H+L), HRP conjugated (1:2000)(Pierce) was diluted 1:2000 in blocking buffer, 100 µl was added to the wells and incubated for an additional 60 minutes. Following three washes, 100 µl of ABTS One Component (Seracare Inc.) substrate was added to the wells and incubated for 30 minutes. The reaction was stopped with the addition of 100 µl of 1% SDS and the optical densities were read at 405 nm. Cutoffs for 80% neutralization titers were determined on each plate. Wells with an $OD_{405}$ less than 20% of the mean value of nine virus only controls, plus one standard deviation, were considered neutralizing.

### 2. eFigures

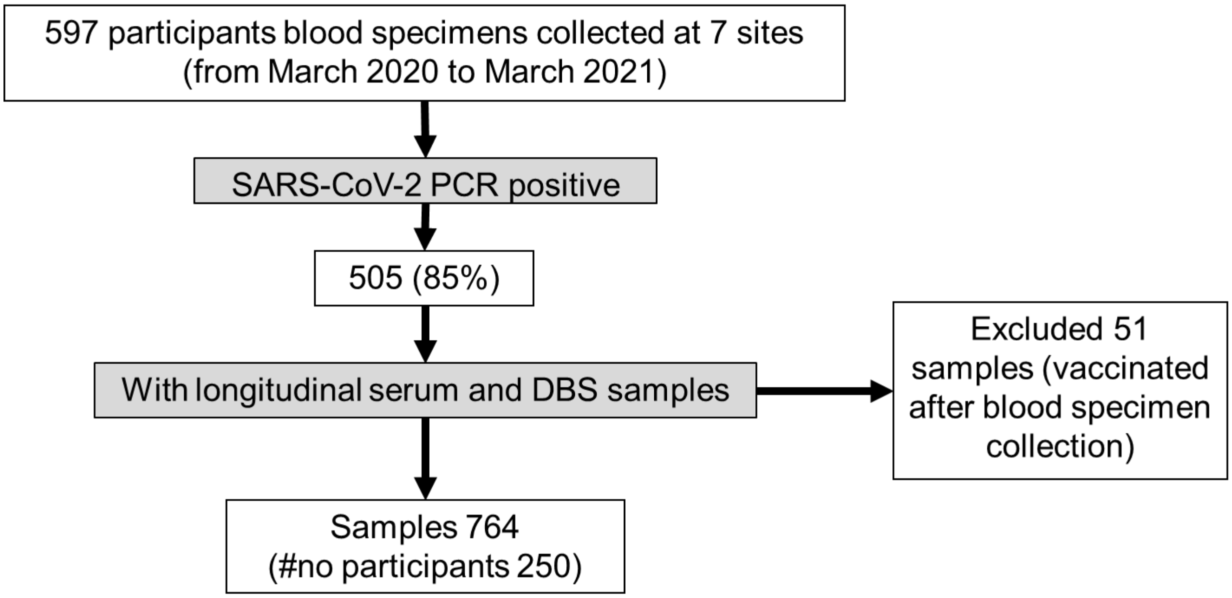


**eFigure1: Flowchart diagram of included participants.**

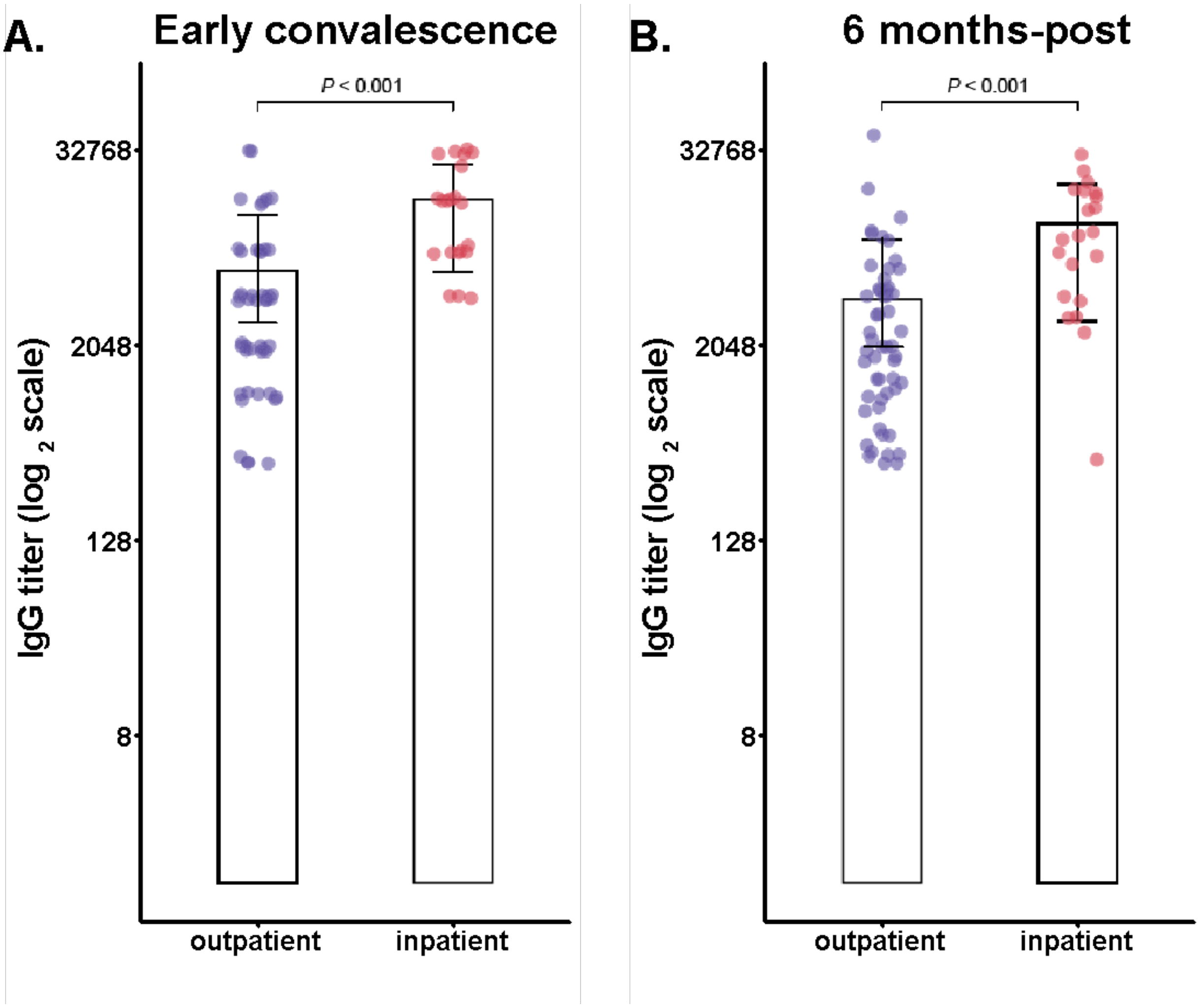


**eFigure 2: The magnitude and durability of anti-SARS-CoV-2 IgG endpoint titers are associated with COVID-19 severity.** Serum samples collected during early convalescence (median 36 dpso) and six months-post (median 188 dpso) from outpatients (n=47) and inpatients (n=21) for serially diluted 2-fold from 1:500 to 1:32,000 to determine endpoint titers, the highest dilution with IgG binding >4774 MFI. Endpoint titers were log-2 transformed and parametric t-tests with Welch's correction applied to determine statistical significance (α=0.05); error bars indicated the geometric mean and 95% confidence intervals.

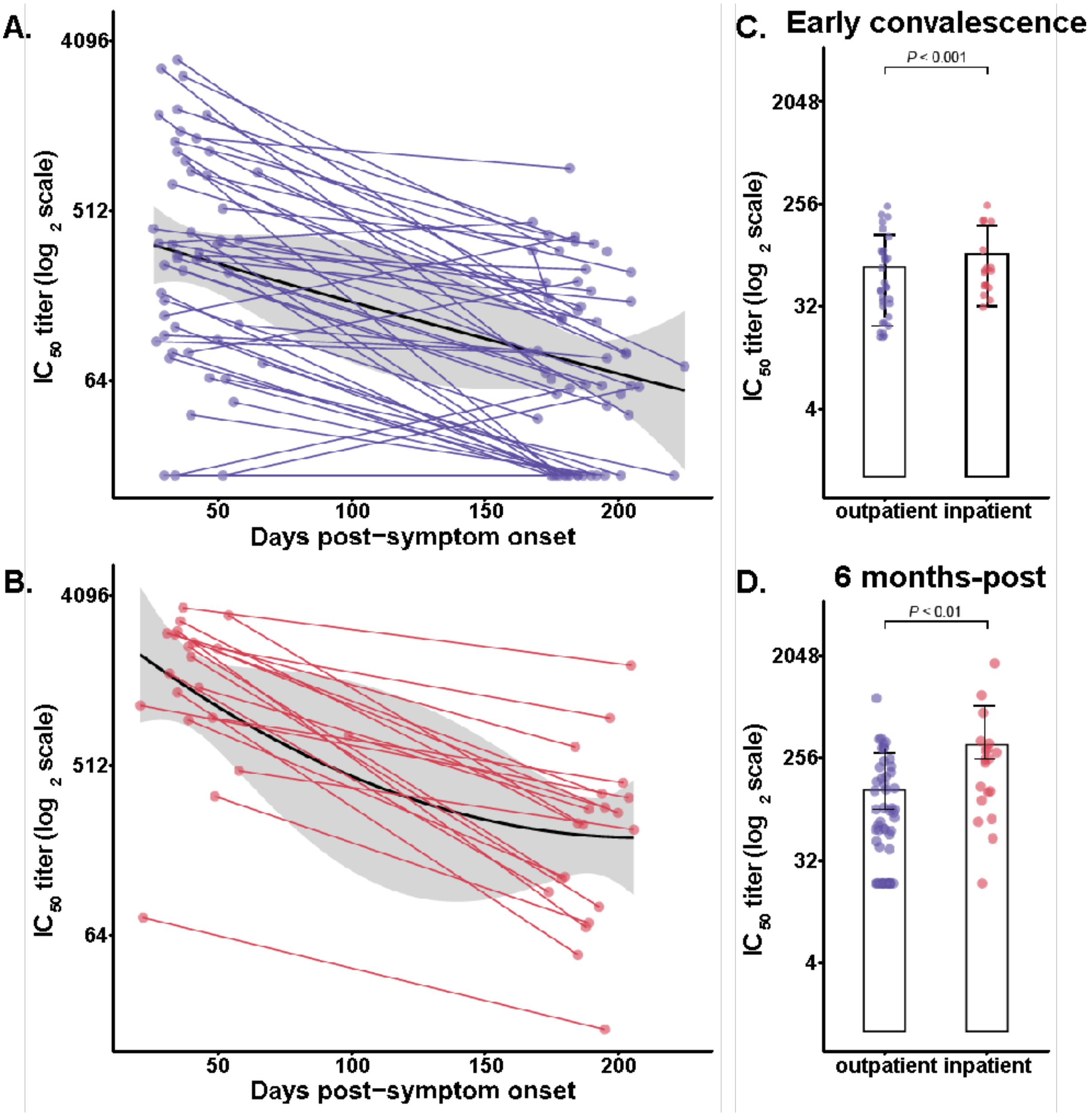


**eFigure 3: Neutralizing antibodies measured by SNT and reported as an IC50 titer.** **(A)** Longitudinal outpatient samples had a half-life of 33 days. **(B)** Neutralizing antibody responses during early convalescence are significantly higher for inpatients. **(C)** Longitudinal inpatient samples had a half-life of 139 days; black lines indicate the second order polynomial curves with 95% CI shaded grey. **(D)** Neutralizing antibody responses six months-post significantly higher for inpatients; error bars indicated geometric mean and 95% CI, statistical significance was determined by parametric t-test with Welch's correction.

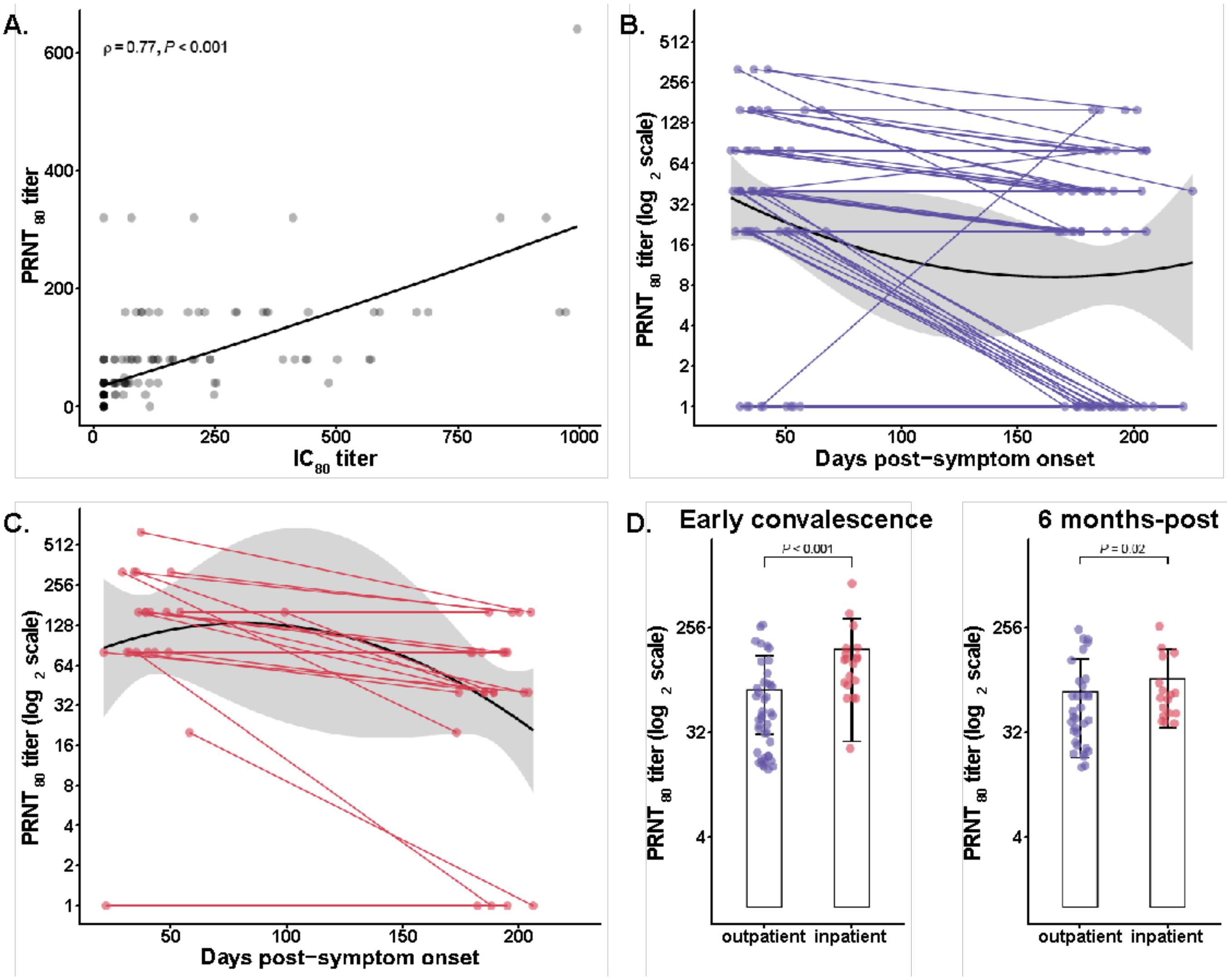


**eFigure 4: Neutralizing antibodies measured by authentic wild-type SARS-CoV-2 plaque reduction neutralization test (VNT).** **(A)** Correlation between SNT $IC_{80}$ and VNT $PRNT_{80}$, Spearman's correlation coefficient is indicated. Longitudinal neutralizing antibody responses of **(B)** outpatients and **(C)** inpatients. **(D)** Neutralizing antibody responses during early convalescence and six months-post were significantly higher for inpatients; error bars indicated geometric mean and 95% CI, statistical significance was determined by non-parametric Mann-Whitney tests.

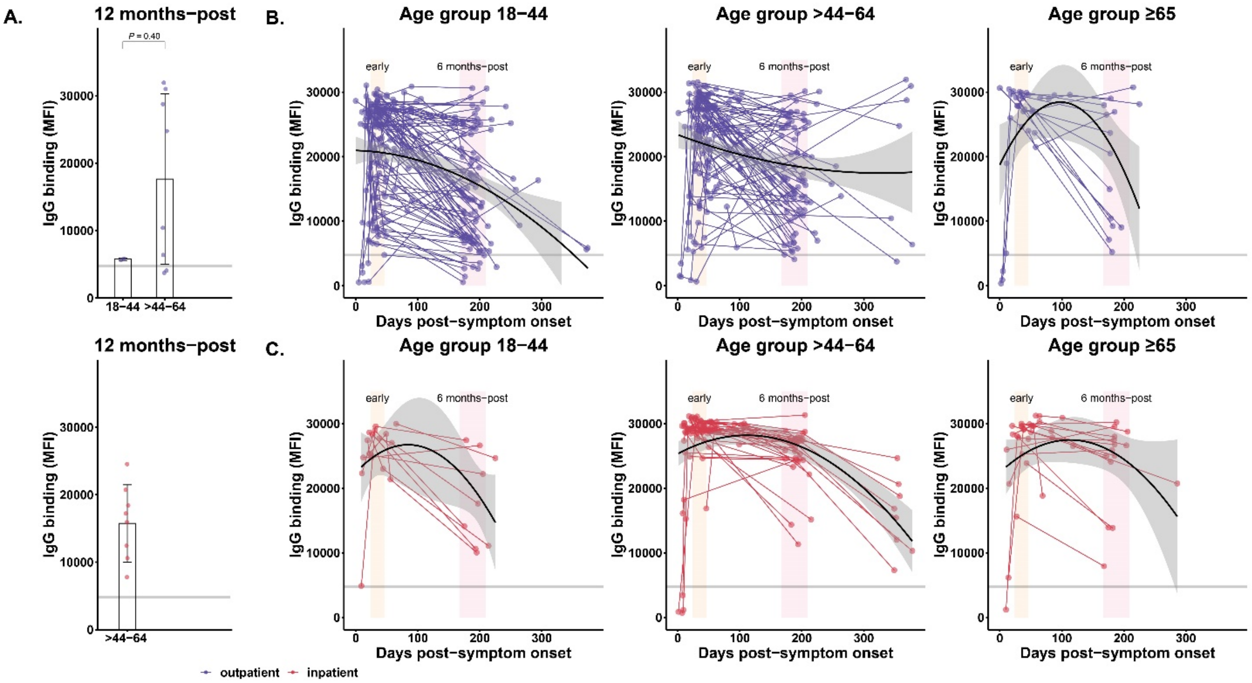


**eFigure 5: The magnitude and six months-post durability of IgG-binding responses are associated with COVID-19 severity and age.** (A) IgG responses twelve months-post compared between outpatient and inpatients. Statistical significance were determined by unpaired t-test with Welch's correction, α = 0.05; error bars indicate the geometric mean and 95% CI. (B) Outpatients stratified by age groups, 18-44 (n=94), >44-64 (n=78), ≥65 (n=14), longitudinal samples are connected by lines, second order polynomial curves and 95% CIs are shaded gray. A horizontal line indicates cutoff for positive/negative IgG; MFI, median fluorescence intensity. Two distinct shaded regions highlighted early convalescence (yellow) and six months-post (pink) windows. (C) Longitudinal inpatient IgG responses stratified by age groups, 18-44 (n=9), >44-64 (n=33), ≥65 (n=16).

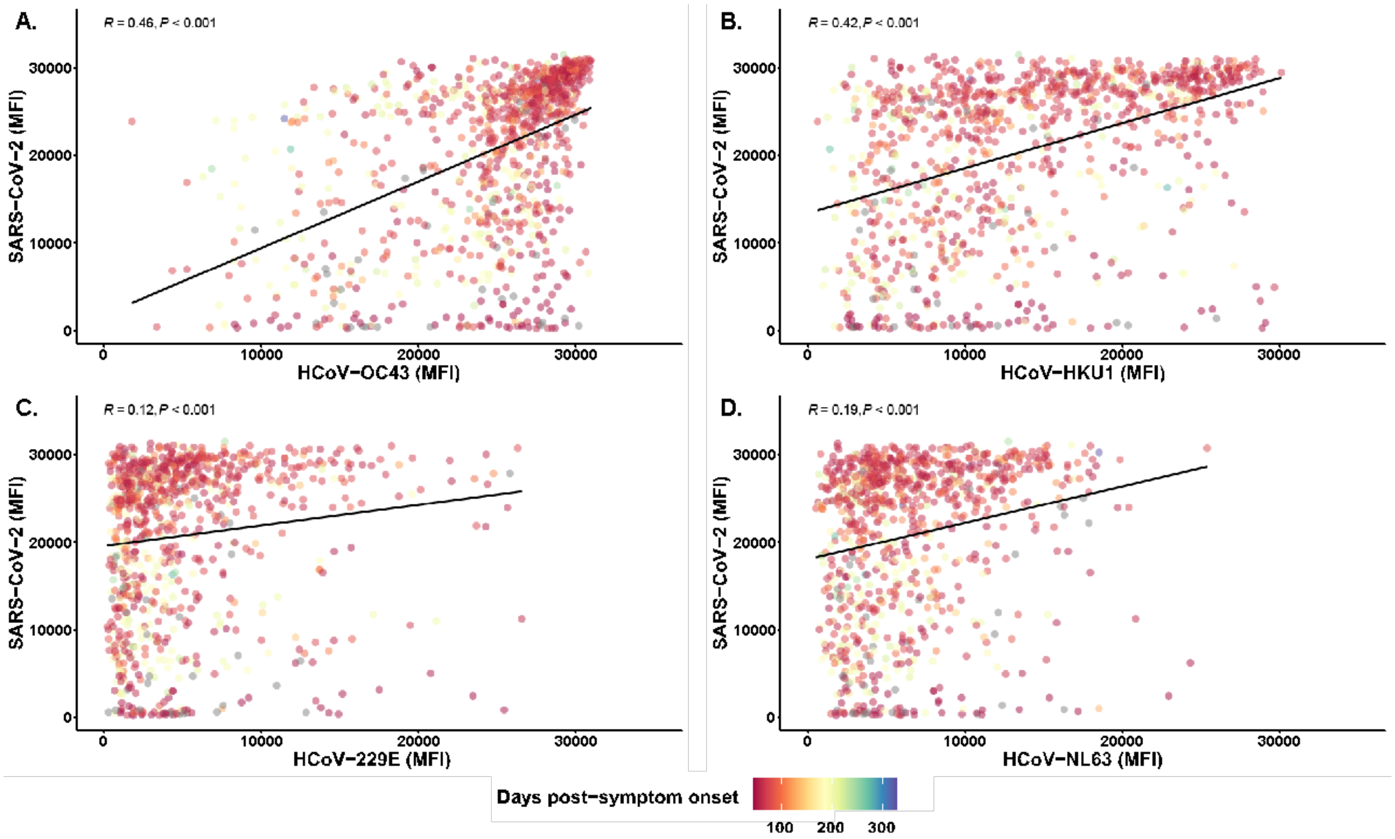


**eFigure 6. The magnitude of seasonal HCoVs antibody responses after SARS-CoV-2 infection**.
Correlation of seasonal HCoVs IgG binding measured against SARS-CoV-2 IgG binding. Each dot represents each sample's IgG level at each timepoint. Dot color depicts dpso from zero to twelve months. (A-B) HCoV-OC43 and HCoV-HKU1 show strong clustering with higher IgG binding MFI levels of SARS-CoV-2 at early time-period post symptom onset. (C-D) Contrastingly higher IgG levels of SARS-CoV-2 were associated with lower IgG levels of HCoV-229E and HCoV-NL63, respectively. The Pearson correlation coefficient was used to measure the strength of the correlation between SARS-CoV-2 and seasonal HCoVs. The correlation coefficient was calculated using Student's t-test, a P-value < 0.05 was considered statistically significant.

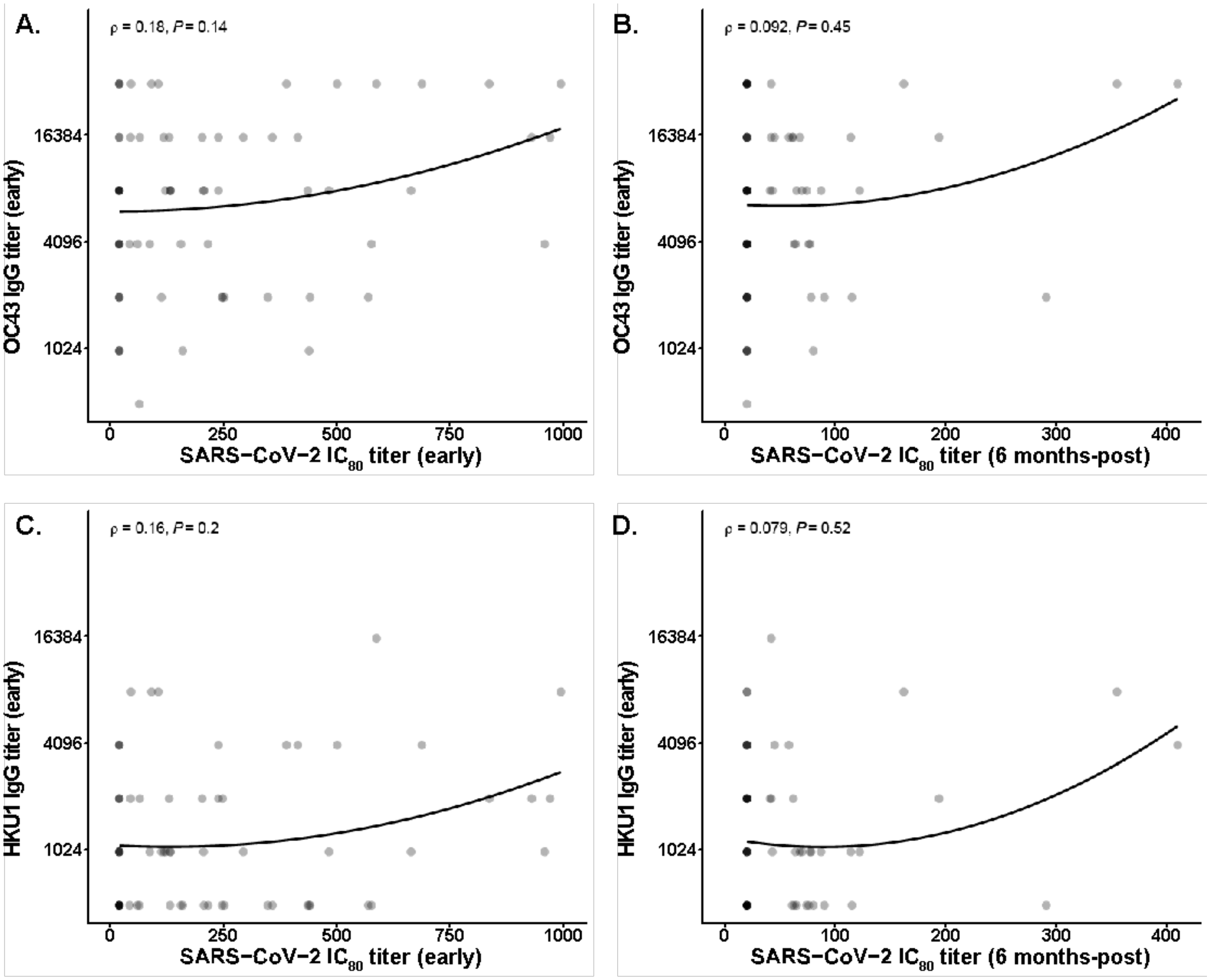


**eFigure 7: HCoV-OC43 and HCoV-HKU1 IgG titers are not associated with SARS-CoV-2 neutralizing antibody responses.** The HCoV-OC43 spike protein IgG titer during early convalescence post-SARS-CoV-2 infection had no significant relationship and low correlation (Spearman's correlation coefficient) with SNT SARS-CoV-2 neutralizing antibody titers during early **(A)** and six months-post **(B)** SARS-CoV-2 infection time periods. Similarly, the HCoV-HKU1 spike protein IgG titer during early convalescence post-SARS-CoV-2 infection had no significant relationship and low correlation (Spearman's correlation coefficient) with SNT SARS-CoV-2 neutralizing antibody titers early **(C)** and six months-post **(D)** SARS-CoV-2 infection time periods.
